# Molecular and spatial epidemiology of HCV among people who inject drugs in Boston, Massachusetts

**DOI:** 10.1101/2022.03.17.22272572

**Authors:** Thomas J. Stopka, Omar Yaghi, Min Li, Elijah Paintsil, Kenneth Chui, David Landy, Robert Heimer

## Abstract

Integration of genetic, social network, and spatial data has the potential to improve understanding of transmission dynamics in established HCV epidemics. Sequence data were analyzed from 63 viremic people who inject drugs recruited in the Boston area through chain referral or time-location sampling. HCV subtype 1a was most prevalent (57.1%), followed by subtype 3a (33.9%). The phylogenetic distances between sequences were no shorter comparing individuals within versus across networks, nor by location or time of first injection. Social and spatial networks, while interesting, may be too ephemeral to inform transmission dynamics when the date and location of infection are indeterminate.

## INTRODUCTION

Although now curable, hepatitis C virus (HCV) infections continue to be problematic. In the United States, HCV mortality has increased in recent years, while mortality for 61 other federally reportable infectious diseases, including HIV, has decreased.^1^ HCV infections have increased by more than 70% among 15-30 year-olds during the past decade, and new infections are largely attributed to injection drug use (IDU).^1,2^ In Massachusetts, over 2,000 new cases attributed to IDU have been reported in those <30 years of age in each of the last seven years.^3^ More information about transmission networks might improve prevention efforts.

The application of phylogenetic and spatial analysis of HCV has identified HCV subtypes among people who inject drugs (PWID) and provided biological evidence of disease clusters and transmission patterns in outbreaks.^4-6^ We aimed to find evidence of disease clusters or transmission patterns among PWID outside of known outbreaks. As part of a study of PWID in the Boston area, we identified a subsample with active HCV infection and sequenced part of their viral genome. We then integrated behavioral, social network, spatial, and HCV phylogenetic data to explore transmission dynamics in established epidemics. We were especially interested to see if we could find evidence of disease clusters or transmission patterns that could identify potential targets, methods, or venues for HCV prevention and treatment interventions.

## METHODS

We employed a cross-sectional design focused on PWID in the Boston area. PWID were eligible if they met the following criteria: (1) age 15-45 years; (2) English or Spanish speaking; (3) evidence of track marks; (4) living with, at risk for, or infected with and subsequently cleared HCV; and (5) ability to provide written informed consent. The study protocol was approved by the [Blinded] Institutional Review Board and the [Blinded] Human Investigation Committee. Consenting individuals completed a survey and were administered the Oraquick® HCV Rapid Antibody test (OraSure Technologies).

Between February and October 2016, recruitment relied on time-location sampling and chain referral in Boston and Cambridge. For the ***time-location sampling*** approach, trained outreach staff recruited participants through partnership with harm reduction programs, shelters, and social service agencies. Staff members at community-based agencies posted study flyers and identified PWID whom they knew were likely to have acquired HCV and would be interested in participating in the study. For the ***chain-referral approach***, enrolled participants (i.e., “seeds”) were asked to refer peers within their networks to the study. In both approaches, research staff described the study to interested recruits, confirmed participant eligibility, provided written materials, and obtained written informed consent. Analysis of recruitment chains was depicted using the StatNet and UserNetR packages in R.

Participants were tested for HCV antibodies and then completed the survey on a tablet, laptop, or desktop computer in our research offices or on tablets or laptops in private spaces provided by community-based partners. Surveys covering demographic, health, and socio-behavioral topics were conducted in English or Spanish and lasted approximately 60 minutes. Participants were informed of HCV test results after completing the survey, and those who tested positive were invited to a separate appointment and were consented to have a blood sample taken. Sample collection continued until 102 individuals were enrolled in the sub-study of HCV-positive participants.

Blood specimens were centrifuged, and the serum was added to RNA*Later*® to preserve the integrity of viral RNA (RNALater; Ambion Inc., Austin, TX). Samples were stored at -80°C. After thawing, viral RNA was purified and a 360-base region in the core gene was amplified and sequenced to identify genotypes and produce phylogenetic trees. We optimally aligned the nucleotide sequences using the CLUSTAL W program and constructed a phylogenetic tree by the Neighbor-Joining method based on Kimura’s two-parameter distances (Mega software, version 7.0). We assessed the reliability of the tree topologies by bootstrapping with 100 replicates, with a bootstrap agreement of ≥70% required to define a phylogenetic cluster. Evolutionary distances were computed using the Maximum Composite Likelihood method.^7^

We compared sequence similarity by recruitment pattern, location of injection initiation, and number of years injecting drugs. We conducted Fisher’s exact tests to assess whether differences in HCV subtypes by recruitment approach were statistically significant. We subsequently ran simple logistic regression analyses to determine whether place of residence at the time of first injection or number of years injecting was significantly associated with HCV subtypes. Place of residence was divided into three categories: Boston, suburbs of Boston (i.e., outside of the city limits and on a Massachusetts Bay Transit Authority (MBTA) line), or beyond the suburbs (not reachable by MBTA line). The number of years injecting was also divided into three categories: 0-5, 6-10, >10 years.

## RESULTS

Among the 102 participants who tested positive for HCV antibodies and provided a subsequent blood sample, 66 were actively infected and sequence data were obtained from 63 of the 66. Out of these 63, 60 completed the survey, and one additional participant completed the survey without providing sequence data. Thus, the sample sizes are 63 for the sequence data and 61 for the survey. The mean age of the 61 respondents was 32±6 years, and males comprised 75% of the sample. Forty (66%) described themselves as White non-Latino, nine as Latino, eight as mixed-race or other, and four as African American. Most were single, with 42 (69%) living alone and 9 (15%) living with a partner. About three-quarters (n=46) reported their sexual orientation as heterosexual, nine as bisexual, and six as gay or lesbian. Three-quarters of the sample reported being homeless and the median monthly income was $500-1000. Ninety percent (n=53) reported having been incarcerated. The mean duration of IDU was 12 years (SD=8) (Table 1).

**Table.**
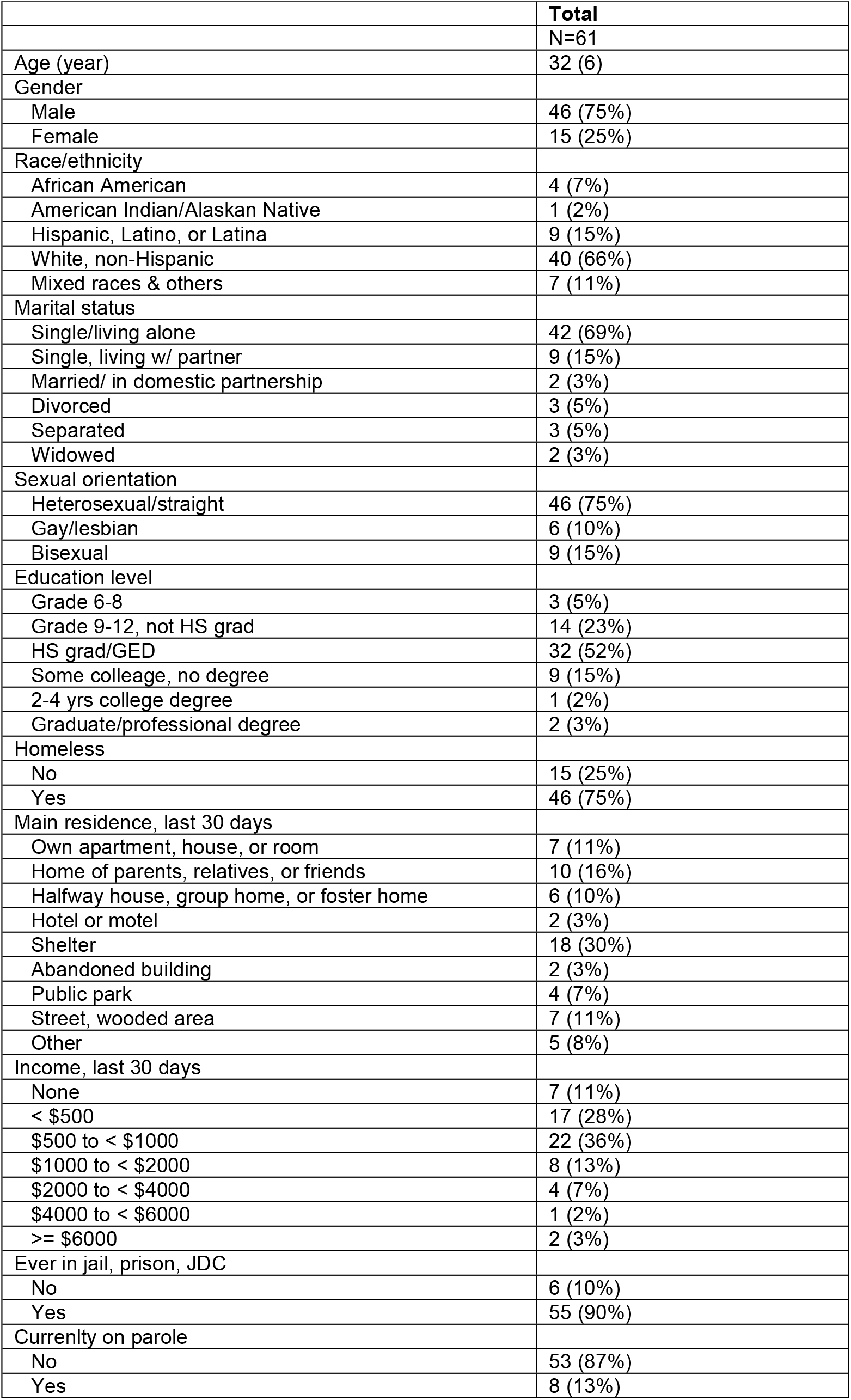
Descriptive statistics: People who inject drugs in the Greater Boston Area of Massachusetts, 2016 (n=61).

### HCV subtype by recruitment pattern

Figure 1A shows the participant network. Thirty-one of the 63 participants were identified through chain referral, indicated by being connected with double-headed arrows, and 32 were identified through time-location sampling, indicated by nodes without connection. Genotype 1a was higher for time-location participants (56.3%; 18/32) than for chain referred participants (41.9%; 13/31; p-value: 0.211); genotype 3a was higher for chain referred participants (45.2%; 14/31) than for time-location participants (22.6%; 7/32; p-value: 0.109). While there was no exclusive clustering of subtypes within the chain referral clusters, three clusters with n>3 only consisted of two subtypes. In addition, in the largest clusters (n=13), seven participants with subtype 3a were connected.

**Figure 1.**
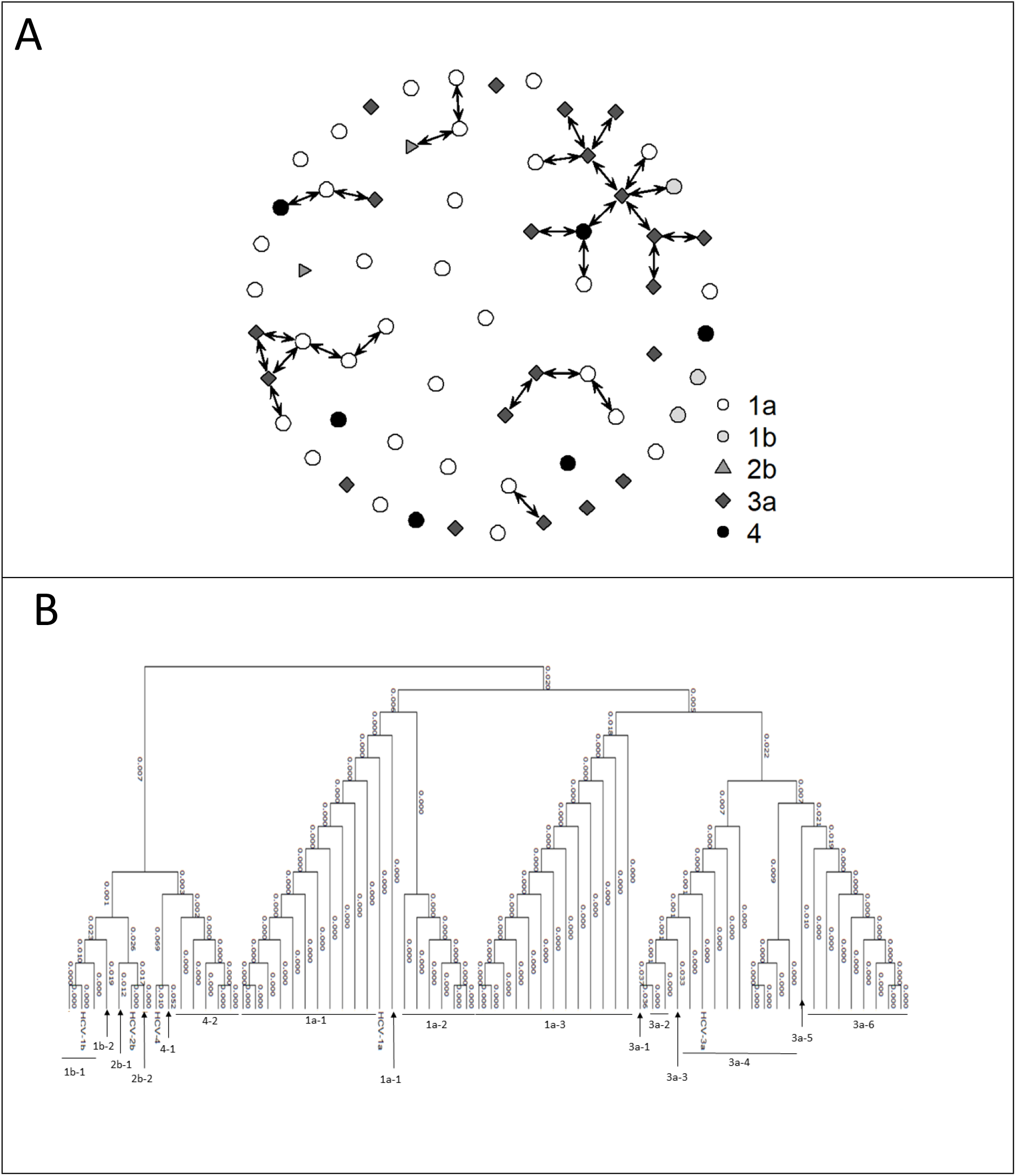
A) Network diagram showing the referral network. The different node symbols represent the five major hepatitis C virus (HCV) subtypes. The double-headed arrows indicate connection by referral (n=31). B) Phylogenetic tree for HCV positive people who inject drugs in Boston, Massachusetts, 2016 (n=63).

### HCV subtype by location of first injection

Among PWID with Subtype 1a, 32.4% (11/34) first injected in Boston, 20.6% (7/34) first injected in Boston suburbs, and 47.1% (16/34) first injected outside of the Boston suburbs. Among PWID with subtype non-1a, 39.1% (9/23) first injected in Boston, 30.4% (7/23) first injected in Boston suburbs, and 30.4% (7/23) first injected outside of the suburbs. Location of the first injection was not associated with HCV subtype, when comparing subtypes from outside of the Greater Boston Area (odds ratio [OR]: 1.87; 95% confidence interval [CI]: 0.54, 6.53) or in Boston suburbs (OR: 0.82; 95% CI: 0.21, 3.26) to subtypes from Boston.

### HCV subtype by number of years injecting

Among PWID with Subtype 1a, 21.9% (7/32) had been injecting for 0-5 years, 31.3% (10/32) had injected for 6-10 years, and 46.9% (15/32) had been injecting for >10 years. Among PWID with Subtype Non-1a, 13.7% (3/22) had been injecting for 0-5 years, 13.7% (3/22) had injected for 6-10 years, and 72.7% (16/22) had been injecting for >10 years. We divided the sample into those infected with genotype 1a versus not 1a, and there was no difference between groups by years of injection experience (OR: 0.97, 95% CI: 0.90, 1.04).

### HCV sequencing results

Sequencing results indicated that the distances between sequences in phylogenetic analyses were no shorter comparing sequence data within versus across networks (Figure 1B). Four of the oldest injectors in the sample (individuals born in 1977 or earlier) were infected with HCV genotypes 1b and 2b. Only one other individual was infected with either of these two subtypes (Figure 1B).

## DISCUSSION

Integrating social network, spatial, and molecular data from established epidemics poses unique challenges. These stem from the lack of defined dates of infection, the durability of current social network connections, the multilevel nature of risk environments,^8,9^ and missing data about the full network structure that is obtained when chain referral relationships are the sole data source on social connections. For this reason, we sought to determine if HCV was more closely related (1) when specimens were obtained from within recruitment chains than those obtained by the less directed recruitment achieved by time-location sampling, and (2) when participants were categorized by time and location that their first exposure to HCV had likely have occurred. For the latter analysis, we made the working hypothesis that HCV infection is an early consequence of initiating (IDU). This hypothesis is consistent with observations of high HCV incidence in the first year of injection.^10,11^

Our findings that HCV subtype 1a was the predominant subtype among PWID in the Boston Area (57.1%), followed by subtype 3a (33.9%), differs from studies elsewhere that focused on HCV among PWID. A study from New York, conducted twenty years earlier than ours, found that the predominant genotypes were 1a and 1b, with 3a comprising only 5% of sequences genotyped.^12^ A study from Baltimore, concurrent with ours, found no evidence of genotype 3a.^13^ Our study population was substantially younger than those in the other two studies, consistent with our aim to recruit a younger sample, but differences in genotype were not associated with age. However, there were two subtypes (1b and 2b) that were restricted to older individuals. It appears, however, that these subtypes have failed to spread among younger members of our study population. Analysis of HCV genotypes from PWID in Vancouver revealed a pattern similar to ours, with nearly half their sample infected with genotype 1a and one-third infected with genotype 3a, but the relevance of this finding is limited by the geographic distance between coasts.^6^

HCV phylogenetic clustering has been found in those recently initiating drug injection. Clustering was quite common in the large HIV-HCV outbreak in Scott County, Indiana. But even there, multiple introductions of HCV were identified. Genotypes 1a and 3b accounted for 72% and 20% of the patient samples, respectively, but additional genotypes had entered the mix, and some people were infected with multiple genotypes.^14^ One study from Melbourne found within-network correlation with HCV genotype, but only in individuals infected with genotype 3 and reporting recent drug injection initiation.^15^ In our study, we did not find any significant associations between HCV subtypes by recruitment types, number of years injecting, nor location of first injection.

Several limitations should be considered in interpreting our findings. First, our sample is small and needs to be expanded. Second, while we aimed to use respondent driven sampling (RDS), we ultimately used a modified RDS approach combined with a time-location sampling approach to recruit our sample during a relatively tight timeframe based on funding requirements. Third, we used self-reported measures from our survey. Some questions we asked referred to events from the distant past (e.g., place where first injected, date of first injection), which could have introduced recall bias. Fourth, our results are not generalizable to the entire Boston Area, nor to other cities. Finally, although we sought to recruit a relatively young sample of PWID, many people had long injecting experiences, so their social networks at the time they were infected with HCV may have been different from their social networks at the time of the survey. In lieu of trying to design studies that collect detailed individual network and phylogenetics to reconstruct patterns of disease transmission, it may be more reasonable to collect information sufficient to populate random mixing and other forms of stochastic models.^16^ Sequencing of infectious disease transmission among PWID may be most effective in the case of disease outbreaks.^17^

## Conclusion

Social networks, while interesting, are too ephemeral to inform transmission dynamics if the date and location of infection are indeterminate. Expanding research efforts to obtain extensive social network data from PWID populations in established epidemics may not be worthwhile or cost-effective given the expense of obtaining full social network data. Alternatives, such as quasi network data obtained through RDS and random mixing models, may be sufficient when modeling transmission networks.

## Data Availability

The data will be uploaded to a public repository upon acceptance of the manuscript.

## Acknowledgments

The authors wish to acknowledge the assistance of the following agencies and individuals who contributed to recruitment and data collection efforts through participant referrals or provision of space for data collection activities: AHOPE, AIDS Action Committee, South Boston Behavioral Health Center, Dr. Laura Grubb, Tufts Medical Center, and St. Anthony’s Shelter. The authors also wish to thank Dr. Nadia Abdala for guidance on HCV specimen sequencing and preparation for phylogenetic analyses.

## Contributions

TS conceptualized the study, oversaw all study activities, wrote the initial draft of the manuscript, and obtained the funding for the study. OY assisted with field operations, data collection, data management, and review of the final manuscript. ML conducted phylogenetic analyses, development of data visualizations, and interpretation of findings. EP oversaw all laboratory analyses and interpretation of phylogenetic analyses. KC conducted statistical analyses and assisted with the development of all figures. DL coordinated field activities, including recruitment, data collection, and data management. RH contributed to study conceptualization, the analytical plan, phylogenetic analyses, and interpretation of findings. All authors contributed to, reviewed, and approved the final version of the manuscript.

## References

1. Ly KN, Hughes EM, Jiles RB, Holmberg SD. Rising Mortality Associated With Hepatitis C Virus in the United States, 2003-2013. Clin Infect Dis. May 15 2016;62(10):1287–1288. doi:10.1093/cid/ciw111

2. Suryaprasad AG, White JZ, Xu F, et al. Emerging epidemic of hepatitis C virus infections among young nonurban persons who inject drugs in the United States, 2006-2012. Clin Infect Dis. Nov 2014;59(10):1411–9. doi:10.1093/cid/ciu643

3. Onofrey S, Aneja J, Haney GA, et al. Underascertainment of acute hepatitis C virus infections in the U.S. surveillance system: a case series and chart review. Ann Intern Med. Aug 18 2015;163(4):254–61. doi:10.7326/m14-2939

4. Mathei C, Van Dooren S, Lemey P, Van Damme P, Buntinx F, Vandamme AM. The epidemic history of hepatitis C among injecting drug users in Flanders, Belgium. J Viral Hepat. Jun 2008;15(6):399–408. doi:10.1111/j.1365-2893.2007.00950.x

5. Caraballo Cortes K, Rosinska M, Janiak M, et al. Next-generation sequencing analysis of a cluster of hepatitis C virus infections in a haematology and oncology center. PLoS One. 2018;13(3):e0194816. doi:10.1371/journal.pone.0194816

6. Jacka B, Applegate T, Krajden M, et al. Phylogenetic clustering of hepatitis C virus among people who inject drugs in Vancouver, Canada. Hepatology. Nov 2014;60(5):1571–1580. doi:10.1002/hep.27310

7. Tamura K, Nei M, Kumar S. Prospects for inferring very large phylogenies by using the neighbor-joining method. Proc Natl Acad Sci U S A. Jul 27 2004;101(30):11030–5. doi:10.1073/pnas.0404206101

8. Rothenberg RB, Long DM, Sterk CE, et al. The Atlanta Urban Networks Study: a blueprint for endemic transmission. AIDS. Sep 29 2000;14(14):2191–200. doi:10.1097/00002030-200009290-00016

9. Rhodes T. Risk environments and drug harms: a social science for harm reduction approach. Int J Drug Policy. May 2009;20(3):193–201. doi:10.1016/j.drugpo.2008.10.003

10. van Beek I, Dwyer R, Dore GJ, Luo K, Kaldor JM. Infection with HIV and hepatitis C virus among injecting drug users in a prevention setting: retrospective cohort study. BMJ. Aug 15 1998;317(7156):433–7. doi:10.1136/bmj.317.7156.433

11. Hagan H, McGough JP, Thiede H, Weiss NS, Hopkins S, Alexander ER. Syringe exchange and risk of infection with hepatitis B and C viruses. Am J Epidemiol. Feb 01 1999;149(3):203–13. doi:10.1093/oxfordjournals.aje.a009792

12. Strasfeld L, Lo Y, Netski D, Thomas DL, Klein RS. The association of hepatitis C prevalence, activity, and genotype with HIV infection in a cohort of New York City drug users. J Acquir Immune Defic Syndr. Jul 01 2003;33(3):356–64. doi:10.1097/00126334-200307010-00010

13. Falade-Nwulia O, Hackman J, Mehta SH, et al. Factors associated with phylogenetic clustering of hepatitis C among people who inject drugs in Baltimore. BMC Infect Dis. Nov 10 2020;20(1):815. doi:10.1186/s12879-020-05546-x

14. Ramachandran S, Thai H, Forbi JC, et al. A large HCV transmission network enabled a fast-growing HIV outbreak in rural Indiana, 2015. EBioMedicine. Nov 2018;37:374–381. doi:10.1016/j.ebiom.2018.10.007

15. Sacks-Davis R, Daraganova G, Aitken C, et al. Hepatitis C virus phylogenetic clustering is associated with the social-injecting network in a cohort of people who inject drugs. PLoS One. 2012;7(10):e47335. doi:10.1371/journal.pone.0047335

16. Kaplan EH. Needles that kill: modeling human immunodeficiency virus transmission via shared drug injection equipment in shooting galleries. Rev Infect Dis. 1989 Mar-Apr 1989;11(2):289–98. doi:10.1093/clinids/11.2.289

17. Tumpney M, John B, Panneer N, et al. Human Immunodeficiency Virus (HIV) Outbreak Investigation Among Persons Who Inject Drugs in Massachusetts Enhanced by HIV Sequence Data. J Infect Dis. 09 02 2020;222(Suppl 5):S259–S267. doi:10.1093/infdis/jiaa053

